# COVID-19 in Scottish care homes: A metapopulation model of spread among residents and staff

**DOI:** 10.1101/2021.08.24.21262524

**Authors:** Matthew Baister, Ewan McTaggart, Paul McMenemy, Itamar Megiddo, Adam Kleczkowski

**Affiliations:** Department of Mathematics & Statistics, University of Strathclyde, Glasgow, UK; Department of Computing Science and Mathematics, University of Stirling, Stirling, UK; Department of Management Science, University of Strathclyde, Glasgow, UK

## Abstract

Care homes in the UK were disproportionately affected by the first wave of the COVID-19 pandemic, accounting for almost half of COVID-19 deaths over the course of the period from 6^th^ March – 15^th^ June 2020. Understanding how infectious diseases establish themselves throughout vulnerable communities is crucial for minimising deaths and lowering the total stress on the National Health Service (NHS Scotland). We model the spread of COVID-19 in the health-board of NHS Lothian, Scotland over the course of the first wave of the pandemic with a compartmental Susceptible - Exposed - Infected reported - Infected unreported - Recovered - Dead (**SEIARD**), metapopulation model. Care home residents, care home workers and the rest of the population are modelled as subpopulations, interacting on a network describing their mixing habits. We explicitly model the outbreak’s reproduction rate and care home visitation level over time for each subpopulation, and execute a data fit and sensitivity analysis, focusing on parameters responsible for intra-subpopulation mixing: staff sharing, staff shift patterns and visitation. The results suggest that hospital discharges were not predominantly responsible for the early outbreak in care homes, and that only a few such cases led to infection seeding in care homes by the 6^th^ of March Sensitivity analysis show the main mode of entry into care homes are infections by staff interacting with the general population. Visitation (before cancellation) and staff sharing were less significant in affecting outbreak size. Our model suggests that focusing on the protection and monitoring of staff, followed by reductions in staff sharing and quick cancellations of visitation can significantly reduce future infection attack rates of COVID-19 in care homes.

## Introduction

The outbreak of the SARS-CoV-2 induced disease (COVID-19) pandemic has had a profound impact, causing 3.7 million deaths by early June 2021 and economic hardship globally [1]. The care home population suffered a disproportionate amount of COVID-19 related deaths. From the week ending 13^th^ March 2020 to the week ending 26^th^ June 2020 (the “first wave”), 54,510 deaths were associated with COVID-19 in the UK, 40% of which were among care home residents [2]. The COVID-19 pandemic has highlighted the vulnerability of care homes, as their resident population is elderly and often suffers from several co-morbidities [3], their systems have not been developed with infection prevention and control (IPC) in mind, and their IPC guidelines have been borrowed from hospitals - a completely different setting [4]. Networks of care homes are an ecosystem connected by staff working across facilities. These connections increase the risk of COVID-19 ingress into care homes, and to protect their vulnerable community, we need to understand the ecosystem dynamics.

Care homes and their residents are enclosed societies, isolated to some extent from the general population. Their connection to broader society primarily consists of interaction with staff and visits from the general population. Care home staff potentially play a vital role in COVID-19 introduction and spread throughout the care home population. Firstly, staff exposure to infection from the general population can establish an outbreak in a home. Secondly, some staff work across multiple homes - a concept we refer to as staff sharing. Staff acting as a bridge between care homes and the general population and staff sharing induces a network, connecting care homes in a given community via their workers. This creates the potential for COVID-19 to spread from one home to another; hence, investigation of this pathway is important. We find it natural to describe this using a heterogeneous patch size metapopulation model framework.

Very few models explore COVID-19 transmission at a community level and explicitly include the unique dynamics in care homes. For example, in [5, 6] agent-based models (ABMs) of single homes are used to investigate the impact of testing strategies on the disease burden. A report by Nyguen et al. [7] uses an ABM to investigate the impact on care home residents of various vaccine coverage, and reducing the weekly testing of staff. However, the models in [6, 7] do have an external force of infection (FOI) from the community, based on prevalence data, representing staff interaction with the community and visitors. These models [5–7] do not assess the relative impact of the different COVID-19 pathways into care homes. Nguyen et al. [8] extend [6, 7], using a hybrid ABM-System Dynamics model, to explore the conditions under which visitation, heterogeneous care homes sizes, and the cohorting of residents impacts COVID-19 outbreak severity.

Rosello et al. [9] model an individual care home with a stochastic compartmental model, using multiple FOI’s to capture COVID-19 pathways, including visitors, hospital discharges, staff working at other homes, and staff infections from the community. They find that importations of infections by staff from the community are the main driver of outbreaks, and importation by visitors or from hospitals is rare, but do not explicitly model disease spread throughout a network of care homes. In [10] individual care homes and the general public are independent, deterministic SEIR models, with a stochastic external FOI connecting the general public to each home. This FOI depends on the prevalence of COVID-19 in the general public, and the size of each home. Transmission rates in homes and in the general public do not vary over time. In [11], two weakly-coupled SEIR sub-models with time-dependent transmission rates define the dynamics; one sub-model describes the general public and one describes all care home residents in Stockholm as a single homogeneous group. Again, a single FOI acts on the residents to capture infections from staff and visitors. The models [10, 11] do not differentiate between, and therefore allow comparison of, the COVID-19 pathways into care homes. Bunnik et al. [12] use a compartmental metapopulation model to explore the trade-offs between increasing protection for a “vulnerable” population and relaxing restrictions for the “non-vulnerable” after the first lockdown in Scotland. They use time-dependent transmission rates with three metapopulation groups; vulnerable, shielders and general public. We extend and apply the methodology of [12] in our model, investigating protection to a vulnerable group (care home residents) in ways other than shielding.

We construct a **SEIARD** compartmental metapopulation model to describe the first wave of COVID-19 in a health board in Scotland. The population is divided into groups of care home residents, staff, and general public. Our care home resident group are not a single homogeneous unit as in [11, 12] but are separate units, creating a refined spatial/geographic structure. These units are not independent as in [10] but are linked by a staff sharing network which, to our knowledge, is unique. We calibrate this model to data from the NHS Lothian Health Board and explore the sensitivity of the results to changes in key parameters. We investigate the importation of infections by staff from the community, visitation, staff sharing, and additionally, we shed light on the exposure of care homes at the beginning of the first wave, e.g., via hospital discharges [13]. The aim is to identify and rank the modes of COVID-19 ingress into and throughout the susceptible care home community in order to improve future pandemic responses. Our model allows for this investigation by coupling the general public and individual care homes with the explicit movement of staff and visitors between the two.

## Materials and methods

### Mathematical model

We develop a deterministic **SEIARD** compartmental metapopulation model with heterogeneous subpopulation sizes. Each subpopulation consists of a host human population, categorised further into six compartments of COVID-19 infection status: Susceptible (**S**), i.e., everyone who is not infected; Exposed (**E**), those exposed to the virus (and infected) but not yet infectious; Infectious and reported (**I**), infectious individuals that have been identified with a positive test; Unreported infectious (**A**), infectious individuals that have not been identified with a positive test; Recovered (**R**), those who had COVID-19 and recovered; and Dead (**D**), those who died from their illness. Symptomatic and asymptomatic individuals are not modelled explicitly; instead, asymptomatic infections contribute towards a reduction in the reporting rates. This model is illustrated in Fig 1 (a).

**Fig 1.**
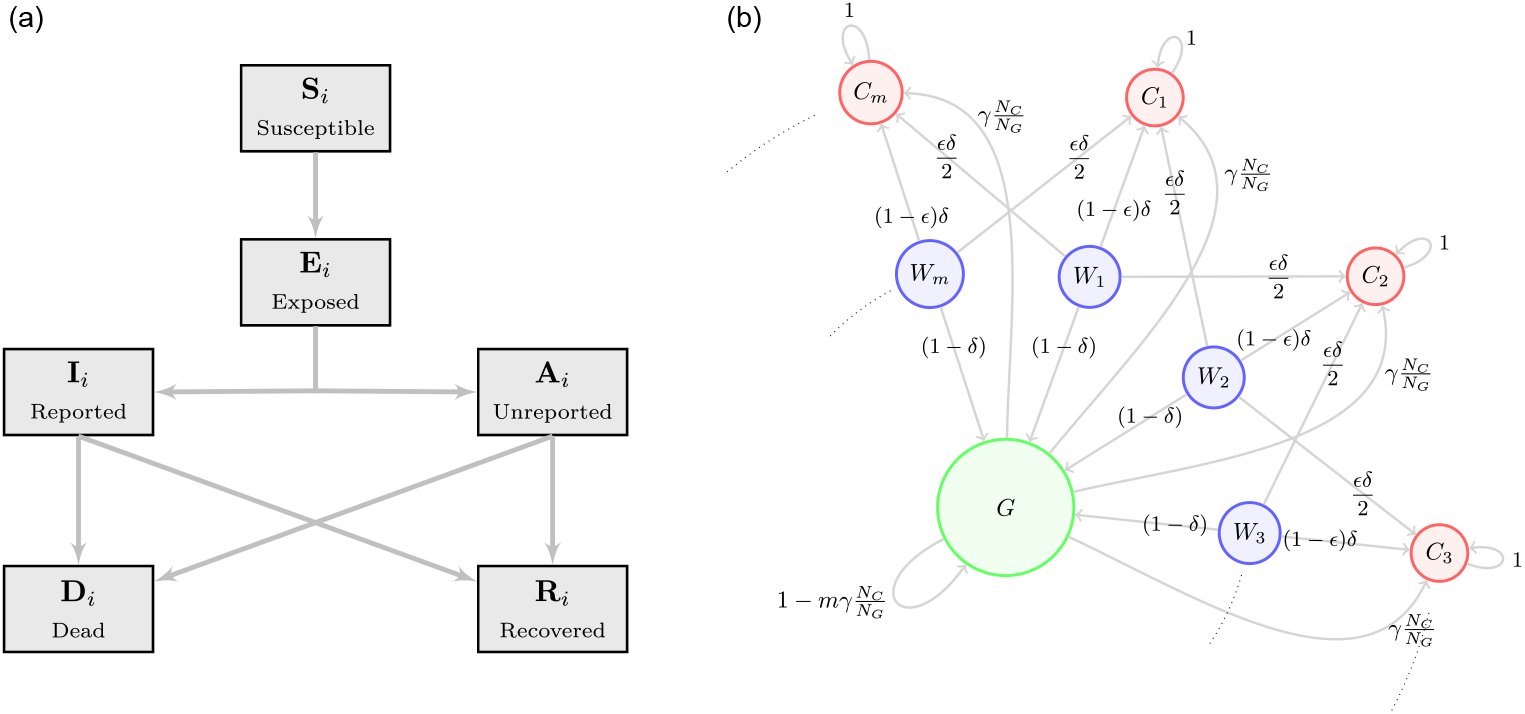
Schematics for the compartmental and metapopulation structure. **(a)**: **SEIARD** compartmental structure of the model; **(b)**: Time-share network of interaction amongst subpopulations. Directed edge weights are *t*_*ik*_, the proportion of people from subpopulation *i* who travel to mix at effective population *k*.

The metapopulation structure represents the population of the NHS Lothian health board in Scotland. We distinguish between care home residents, care home workers and the general population, modelling the *m* = 109 care homes for older adults in NHS Lothian [14]. The *j*^*th*^ home has a resident subpopulation, *C*_*j*_, with a corresponding care home worker subpopulation, *W*_*j*_. The general population is encapsulated by the subpopulation *G*. Each care home includes the same number of residents, a simplifying assumption made due to lack of publicly available data on care home sizes in Lothian. We also assume the worker subpopulations are the same size as residents’ [15].

Each node of the network, *i* ∈ *X* := {*C*_1_, *C*_2_, …, *C*_*m*_, *W*_1_, *W*_2_, …, *W*_*m*_, *G*} with |*X*| = *n*, is described in terms of the **SEIARD** compartmental model with equations:

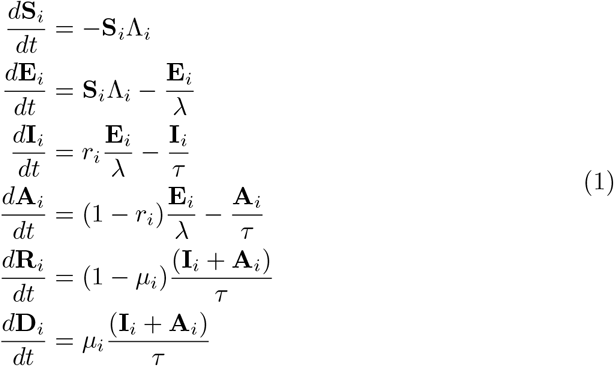

Susceptibles in subpopulation *i* (**S**_*i*_), are infected with a FOI Λ_*i*_, and moved to the exposed class (**E**_*i*_). After a non-infectious latent period of *λ* days, they become infectious, testing positive at a reporting rate of *r*_*i*_. These identified infections move to the class **I**_*i*_. Hence, any unidentified infections, **A**_*i*_, occur at rate 1 - *r*_*i*_. After *τ* infectious days, a person either recovers or dies at the rate *μ*_*i*_. For simplicity, and considering the short span of time the model is designed to describe, non-COVID related deaths are not considered. For similar reasons, we do not include a birth rate or admission of new residents to care homes from the general population.

We assume a constant reporting rate for care home residents 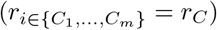, workers 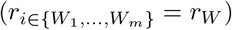, and the general public (*r*_*G*_). The parameters *τ* and *λ* describe the infectious period and latency period, respectively, and are assumed to be the same across all subpopulations. Mortality rates, *μ*_*i*_, vary by subpopulation, reflecting the positive association of serious outcomes of COVID-19 with age [16]. As we are modelling over a period of 4 months (approx. first wave), and immunity after COVID-19 infection lasts as long as 5 months [17] [18], we do not consider a transition from Recovered to Susceptible.

We model visitation to each care home by multiplying the proportion of the population, *N*_*C*_*/N*_*G*_, that visit the care home, and their length of stay, *γ*(*t*). The proportion remains constant over time while *γ*(*t*) varies over time. Up until 13^th^ March 2020, each resident has one visitor per day [6]. Then *γ*(*t*) drops to 0, reflecting the policy change to essential visitation only [14, 19]. *γ*(*t*) is described by the function logi(Ω_*x*_), with 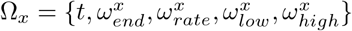, defined below:

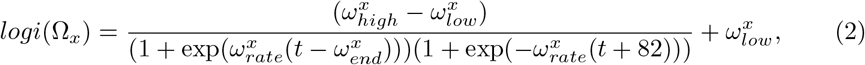

with the shape of a sigmoidal logistic function. The value of 82 is used in the function so that when *t* = 0, *logi*(Ω_*x*_) = *ω*^*high*^. The function drops from *ω*^*high*^ to *ω*^*low*^ at a time controlled by 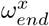, such that when 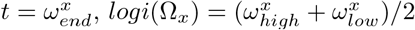. The *ω*_*rate*_ parameter changes the gradient of the descent at *t* = *ω*_*end*_.

Thus, visitation rate is described by 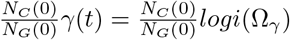. Given that visitation drops to 0 in the first 2 weeks of the simulation, the changes in population size over that time is negligible, hence we can keep the proportion of the population constant and control visitation by solely changing *γ*(*t*).

A constant proportion of workers, *δ*, spend their time at care homes. With *δ* = 0.5, half of a worker compartment, *W*_*i*_, are at care homes, *C*_*i*_, over the course of a day. The proportion of workers not at care homes, 1 − *δ*, reside in the general population, *G*. During the first wave of COVID-19 in Scotland, there was both intra-organizational staff sharing between homes (i.e., staff who work at multiple homes belonging to the same care provider), as well as inter-organizational staff sharing (use of bank or agency staff) [20, 21]. Therefore, a constant proportion of each homes’ assigned workers, *ε*, are exchanged between homes. We refer to this as *staff sharing*. We have made the simplifying assumption that the staff sharing network has a topology of a circle, whereby the shared staff for home *j* are split evenly between homes *j* − 1 and *j* + 1. We assume care home residents do not leave their homes.

Interaction across subpopulations is heterogeneous and is described in terms of time-sharing, determining proportions of subpopulations mixing in groups with each other. In the *i*^*th*^ subpopulation there are *N*_*i*_(*t*) = **S**_*i*_(*t*) + **E**_*i*_(*t*) + **I**_*i*_(*t*) + **A**_*i*_(*t*) + **R**_*i*_(*t*) active individuals who can mix with others. The proportion from subpopulation *i* who travel to, and mix with, subpopulation *k* is *t*_*ik*_. The effective population size of subpopulation *k*, given that others have travelled to it and some people from *k* have left, is 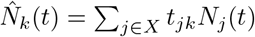. We assume these effective populations 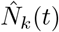 are well mixed, so people who travel to each population can meet all others there. There are two types of effective populations; the care homes and the general population. Care home *j*, comprises 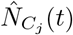 people: its residents, its working staff, staff from other care homes, and visitors. The general population consists of 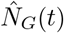 people; this includes all the staff not at work and the non-visiting general population.

Our specific time-share assumptions are represented visually as a directed, weighted network in Fig 1 (b). The corresponding weighted adjacency matrix, the travel/time-share matrix, is ***T*** ∈ ℝ^*n*×*n*^, whose [*i, j*]^*th*^ element is *t*_*ij*_. The rows and columns of ***T*** are in the order of {*C*_1_, *C*_2_, …, *C*_*m*_, *W*_1_, *W*_2_, …, *W*_*m*_, *G*}. ***T*** consists of the partitions {*T*_*CC*_, *T*_*CW*_, *T*_*WC*_, *T*_*WW*_, *T*_*CG*_, *T*_*WG*_, *T*_*GC*_, *T*_*GW*_}. To clarify notation: matrix ***I***_*m*_ indicates the identity matrix of dimension *m*, matrix [*a*]_*m*×*m*_ indicates a matrix of dimension *m* × *m* with all entries *a*. Hence ***T*** and the subsequent sub-matrices are as follows:

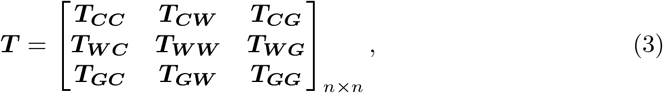

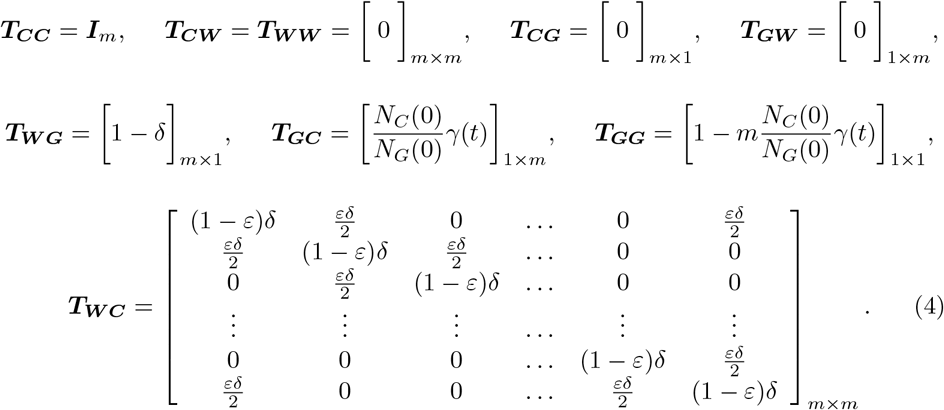

Disease transmission in the model is assumed to be frequency-dependent. The FOI integrates which infections occur to whom, from whom and where the infection takes place, as in [22, 23]. The FOI acting on subpopulation *i*, Λ_*i*_ (see Equation 5), accounts for the different groups’ people from *i* mix in over a day and who they encounter. It is most easily understood by considering Λ_*i*_**S**_*i*_:

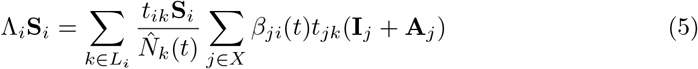

The set of effective populations that subpopulation *i* travels to is *L*_*i*_, consistent with the non-zero elements in the *i*_*th*_ row of the travel matrix ***T***. At effective population *k*, there is *t*_*ik*_**S**_*i*_ susceptible individuals from *i*. At *k* there will also be *t*_*jk*_(**I**_*j*_ + **A**_*j*_) infectious people from *j* who have travelled to *k*. The transmission rate between subpopulation *j* and *i* is *β*_*ji*_(*t*). Therefore,

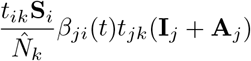

is the number of new daily infections in *i* caused by people from *j* at the effective population *k*.

The transmission rates *β*_*ji*_(*t*) allow us to represent heterogeneous interaction patterns of individuals between and within different subpopulations. They incorporate the transmission dynamics of COVID-19 changing over time and location, for example, through lockdowns or other changes in behaviour [22]. We write 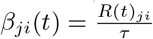, describing the transmission rate *β*_*ji*_(*t*) between subpopulations *j* and *i*, with the reproduction rate, *R*(*t*)_*ji*_, divided by the infectious period, *τ*. The contact rate and infection probability between subpopulations *i* and *j* is captured by *R*(*t*)_*ji*_. We assume only the transmission rates between and within the subpopulation types (residents *C*, workers *W*, general public *G*) differ. Therefore, the transmission rates are arranged in a symmetric partitioned matrix ***β*** ∈ ℝ^*n*×*n*^ whose [*j, i*]^*th*^ element is *β*_*ji*_(*t*). The rows and columns of ***β*** are in the order of {*C*_1_, *C*_2_, …, *C*_*m*_, *W*_1_, *W*_2_, …, *W*_*m*_, *G*}. ***β*** contains block sub-matrices;

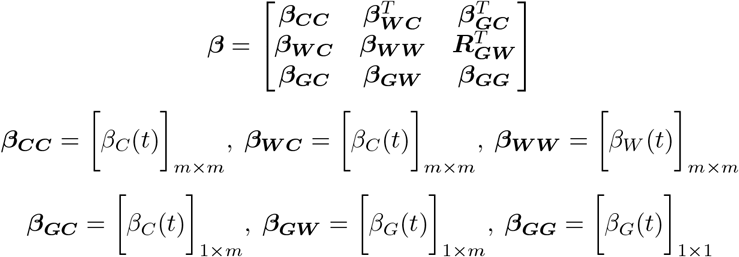

The matrix notation above is the same as for the travel matrix ***T***. For simplicity, we have assumed that the resident-resident, worker-resident, and general population-resident transmission rates are equal. Similarly, we assume the general population-worker and general population-general population transmission rates are the same. The transmission rates are described by:

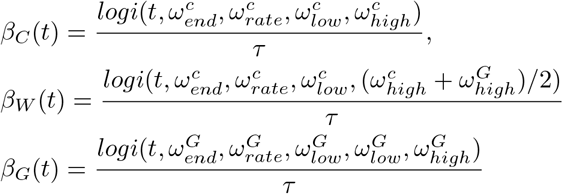

Where the logi function, Equation 2, models the reproduction rate. To simplify and to reduce the number of parameters, we relate the reproduction rate for workers in terms of the residents and general population. As care home workers balance their time between care homes and the general population, we assume the workers pre-lockown maximum reproduction rate is the average of the care homes and general populations, 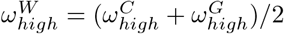. We assume the reproduction rate for workers and residents drops at the same time, and to the same value.

### Model calibration process

We used data from the network of care homes in NHS Lothian [14] complemented by Public Health Scotland Open Data, breaking down COVID-19 cases and deaths per health board [24, 25], to inform and calibrate our model. Parameters were found using a mixture of methods, as indicated in Table 1, including literature search, sensitivity of results, and rigorous fit based on minimising the sum of squares of residuals.

**Table 1.** Parameter definitions, alongside their base case values and source.

| Parameter | Description | Value | Source |
| --- | --- | --- | --- |
| $\varepsilon$ | Staff sharing | 0.4 | Data fit |
| $\delta$ | Proportion of workers at care home | 0.5 | Assumption<sup>a</sup> |
| $r_{i \in \{C_1, \dots, C_m\}} = r_C$ | Reporting rate for residents | 0.53 | Estimated [14, 28–30] |
| $r_{i \in \{W_1, \dots, W_m\}} = r_W$ | Reporting rate for workers | 0.52 | Estimated [14, 24, 27, 30, 31] |
| $r_G$ | Reporting rate for general public | 0.077 | Estimated [24, 27, 31] |
| $\mu_{i \in \{C_1, \dots, C_m\}} = \mu_C$ | Death rate for residents | 0.25 | Estimated [14, 28–30] |
| $\mu_{i \in \{G, W_1, \dots, W_m\}} = \mu_G$ | Death rate for general public (and workers) | 0.017 | Estimated [24, 25, 27, 31] |
| $\tau$ | Infectious period | 7 days | [32] |
| $\lambda$ | Latent period | 5.8 days | [33] |
| $m$ | Number of care homes | 109 | [14] |
| $N(0) = \sum_i N_i(0)$ | Total initial population | 907,580 | [27] |
| $N_{C_i}(0)$ | Initial resident subpopulation size | 48 | [14] |
| $N_{W_i}(0)$ | Initial worker subpopulation size | 48 | [15] |
| $N_G(0)$ | Initial general public subpopulation size | 897,116 | Estimated [14, 15, 27] |
| $\omega_{\text{end}}^C$ | Timing of $R_t$ descent for residents and workers | 42 days | Manually fitted <sup>b</sup> |
| $\omega_{\text{rate}}^C$ | Rate of descent of $R_t$ for residents and workers | 0.5 | Assumption<sup>c</sup> |
| $\omega_{\text{low}}^C$ | Post-descent $R_t$ for residents and workers | 0.6 | Assumption<sup>d</sup> |
| $\omega_{\text{high}}^C$ | Pre-descent $R_t$ for residents | 4.7 | Data fit |
| $\omega_{\text{end}}^G$ | Timing of $R_t$ descent for general population | 22 days | Manually fitted <sup>b</sup> |
| $\omega_{\text{rate}}^G$ | Rate of descent of $R_t$ for general population | 0.5 | Assumption<sup>c</sup> |
| $\omega_{\text{low}}^G$ | Post-descent $R_t$ for general population | 0.6 | [34] |
| $\omega_{\text{high}}^G$ | Pre-descent $R_t$ for the general population | 4.1 | Data fit |
| $\omega_{\text{end}}^\gamma$ | Timing of descent for visitation | 10 days | [14, 19] |
| $\omega_{\text{rate}}^\gamma$ | Rate of descent of visitation | 3 | [14, 19] |
| $\omega_{\text{low}}^\gamma$ | Post-descent value for visitation | 0 | Assumption<sup>e</sup> |
| $\omega_{\text{high}}^\gamma$ | Pre-descent $R_t$ for visitation | 0.083 | Data fit |
| $H_{\text{seeded}}$ | Number of homes seeded | 4 | Data fit |
| $E_G(0)$ | Initial general population infections | 120 | Data fit |

<sup>a</sup> We assume workers spend half day at work, other half mixing in general population. Alternatively, workers do 12hr shifts. Units are given where appropriate in the Value column.
<sup>b</sup> Manually set by matching the model output to the infection peak dates in the NHS Lothian data [14, 24].
<sup>c</sup> Initial model exploration indicated that higher rates (steeper drops in $R_t$ ) resulted in infection peaks (in general public and residents) falling too quickly compared to the NHS Lothian data [14, 24]. The value of 0.5 corresponds to a descent of $\sim 2$ weeks.
<sup>d</sup> We assume the reproductive rate for every sub-population drops to the Scottish government’s [34] estimated $R_t$ after lockdown (so $\omega_{\text{low}}^C = \omega_{\text{low}}^G$ ).
<sup>e</sup> Equals 0 to reflect the policy change to essential visitation only [14, 19], and to avoid the complication of modelling end-of-life visitation.

#### Data

NHS Lothian is the second-largest health board in Scotland [26], providing public health services to an estimated 907,580 people (2019 mid-year population estimate [27]). The daily confirmed positive tests of COVID-19 cases reported across the entire health board were taken from the Public Health Scotland Open Data [24]. This data does not delineate which cases occurred in care homes, and thus, we retrieved the subset of cases in care homes from Burton et al. [14], which reports a 7-day average of confirmed cases in care home residents. Weekly COVID-19 deaths at the NHS Lothian health board level come from National Records Scotland [25]. Care home resident deaths are a subset of these and are published in [14]. Both death data are weekly counts of registered deaths where COVID-19 is mentioned on the death certificate (either as the underlying cause or as a contributory factor) [25].

#### Parameters set using evidence and assumptions

In this section, we describe our assumptions on parameters, parameters estimated from the literature, and parameters manually calibrated to the data.

A Scottish population study between 10^th^ April to 15^th^ June [31] estimated a combined adjusted seroprevalence across their study period (first wave = 10^th^ April to 15^th^ June) of 4.3% (95% CI 4.2%-4.5%). As of the week beginning 15^th^ June 2020, there had been 18,077 positive tests [24], which as a percentage of Scotland’s population (2019 census [27]) is ∼ 0.33%. We use this information to assume a constant reporting rate in the first wave for the general public of *r*_*G*_ = 0.33*/*4.3 ∼ 0.077.

In Scotland, the policy from the start of March to 16^th^ April 2020 was to test only the first few symptomatic care home residents, and afterwards, was to test all symptomatic residents [14]. Assuming when there is an outbreak in a home, 40% of the residents end up infected (40% incidence) [28, 29]. Given 48 residents per care home, until 16^th^ of April we assume a reporting rate of (a few tested)/(total infected) = 3/(0.4 × 48) = 5/32. After 16^th^ April, we assume all the symptomatic cases are reported, giving a reporting rate of 4/5 (an estimated symptomatic proportion of COVID-19 cases in long term aged care is 80% [30]). Between the start of our simulation (6^th^ March 2020) and 16^th^ April 2020 is a time difference of 42 days, and between 17^th^ April 2020 and the end of our simulation period (15^th^ June 2020) is a time difference of 60 days. Therefore, for 42 days, we assume a reporting rate of 5/32, and for 60 days, it is 4/5. The weighted average and constant CH reporting rate over the simulation period is *r*_*C*_= (5/32)(42/102) + (4/5)(60/102) ∼ 0.53.

Until the 17^th^ of April, we assume the staff reporting rate was the same as the general public (0.077). From then on, we assume the care home testing policy change (on the 17^th^ of April) extended to their staff [14], and the reported percentage of cases was 83% (the symptomatic proportion [30]). Our weighted average and constant staff reporting rate over the simulation period is *r*_*S*_ = 0.077×(42/102)+ 0.8360/102 ∼ 0.52.

There are two constant death rates in our model: a resident death rate (*μ*_*C*_) and a general population death rate (*μ*_*G*_). We assume care home staff have the same death rate as the general population. There were ∼ 899 positive tests and 423 deaths in NHS Lothian care home residents over the study period. Using our resident reporting rate, we estimate there were 899.1*/*0.53 ∼ 1697 total residents infected with COVID-19 over the study period. Therefore, we estimate a resident death rate of *μ*_*C*_ = 423*/*1697 ∼ 0.25. Similarly, there were 3123 total positive tests and 709 deaths over the study period in NHS Lothian overall. Using our general reporting rate, *r*_*G*_, we estimate a general population death rate of *μ*_*G*_ = 709*/*(3123*/*0.077) ∼ 0.017.

Under our parameterisation, the timing of the drop in reproductive rates for care home residents 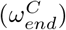 and for the general population 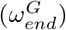 control the timing of peak infections in each respective population, independent of all other parameter values. This is linked to the reproductive rate function (*logi*) at an inflection point at 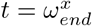 (where *logi*(Ω_*x*_) takes the value of 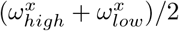. Therefore, we manually set these parameters 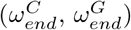 by matching the model output to the infection peak dates in the NHS Lothian data [14, 24].

The 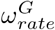 and 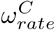 parameters were assumed to be 0.5. Initial model exploration indicated that higher rates (steeper drops in *R*_*t*_) resulted in infection peaks (in general public and residents) falling too quickly compared to the NHS Lothian data [14, 24]. From sensitivity analysis, we found that changing the values of these parameters does not affect the disease dynamics (in terms of total infections/deaths). The *ω*_*rate*_ parameter controls the steepness of the descent from *ω*_*high*_ to *ω*_*low*_, however the timing of the start of the descent changes to almost cancel out the effect of changing the steepness of this drop. The value of 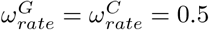, corresponds to a descent of ∼ 2 weeks. For 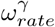 we chose the value of 3 to follow the rapid visitation policy changes in care homes [14, 19].

The 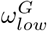 and 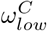 values were set to 0.6, the estimated *R*_*t*_ after the first wave in Scotland [34]. Due to the uncertainty in the timing of the drop and the *R*_*t*_ peak value, we did not use this source for the 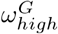 and 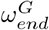 parameters.

We have made the simplifying assumption of 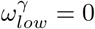, to avoid the complications of modelling end-of-life visitation in care homes.

Closed environments are conducive to COVID-19 transmission and superspreading events [35], therefore we assume pre-lockdown transmission rates within care homes are not less than the general populations, 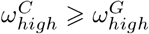.

Care homes operate with staff under differing working hour schedules. This includes care homes having a day and night shift (*δ* = 0.5), three 8 hour shifts (*δ* = 0.33) or an uneven distribution of staff spread throughout the day. For simplicity, we assume that all homes operate under two 12 hour shifts per day, i.e., *δ* = 0.5. Other shifts are explored in the sensitivity analysis.

We make a number of assumptions about the population initially infected. Workers were not initially infected in the model. In the general population, we assume an equal amount of exposed and infected individuals (with and without symptoms) i.e., *E*_*G*_(0) = *I*_*G*_(0) + *A*_*G*_(0). 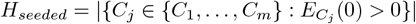 care homes were seeded with infections, representing introductions such as hospital discharges. To account for the delay in infections at the start of the pandemic in care homes compared to the general population, as seen in the data Fig 2, we assume for all 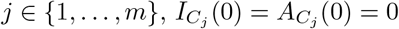. We seeded the homes so that initially infected homes lay equally spaced on the circle sharing structure (see Fig 1 (b)). If a home is seeded then we assume 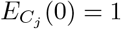, and if not, 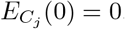.

**Fig 2.**
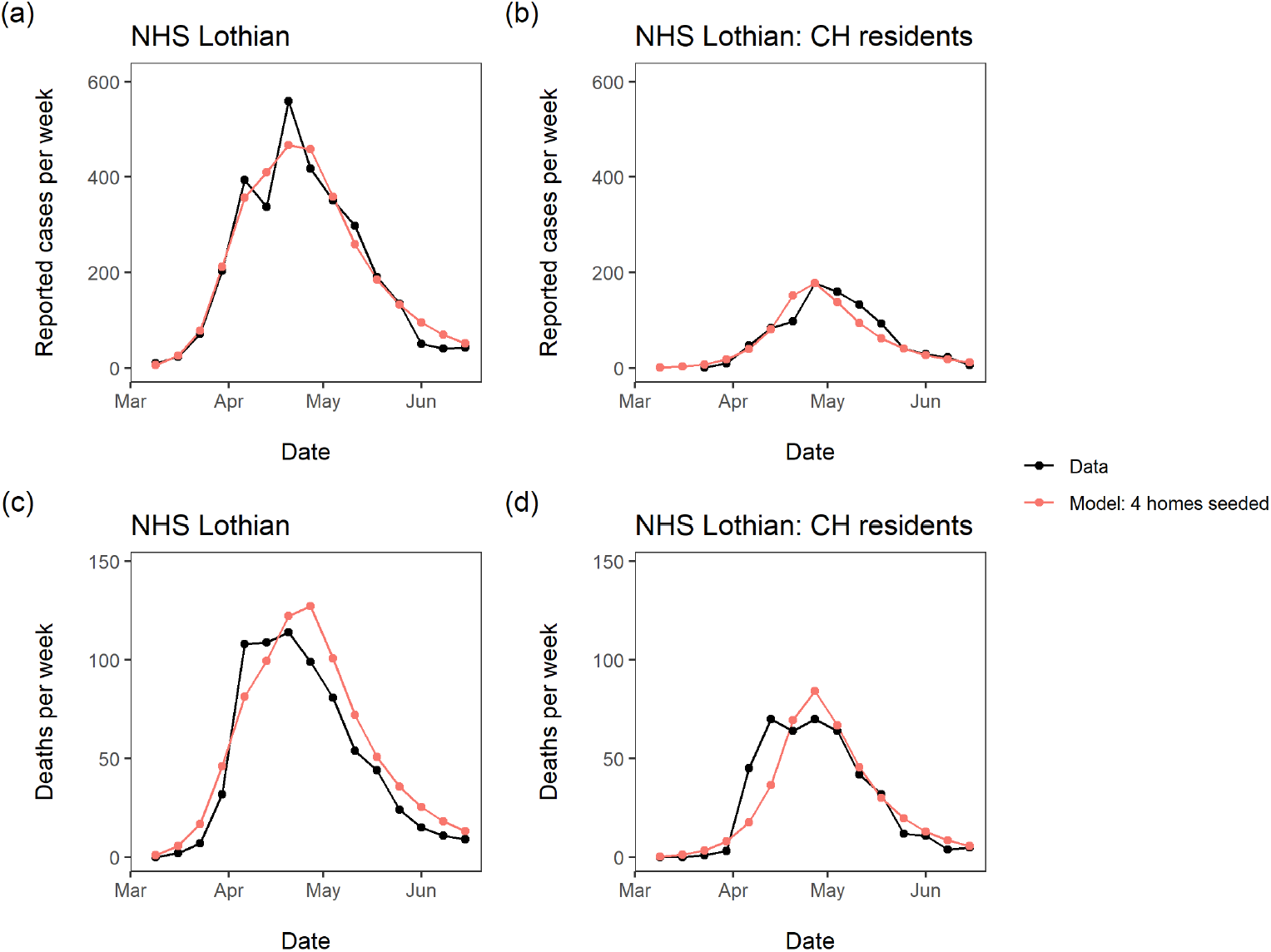
Surveillance data and fitted model. Data used for fitting are black lines, and model solution with parameter values in Table 1 are red lines. (a) reported cases per week for all NHS Lothian inhabitants (care home residents, workers and the general population); (b) reported cases per week in NHS Lothian care home residents; (c) deaths per week for all NHS Lothian inhabitants (care home residents, workers and the general population); (d) deaths per week in NHS Lothian care home residents.

#### Data fit

While some parameter values can be found based on the external data and literature, as shown in the previous section, other parameters were estimated using a formal fit to the cases and deaths data for the Lothian NHS health board (Table 1). These parameters were free to vary subject to constraints based on a combination of assumptions and information from the literature. We used the method of least-squares, aggregating the error of model output against the four data sets for NHS Lothian cases and deaths and choosing the parameter set which minimised this error. The data for NHS Lothian population cases and care home cases were in the form of daily and seven day averages respectively. The death data for both the NHS Lothian population and care homes were in weekly counts. To make the fitting consistent, we transformed the daily and seven day average data into weekly data for conformity (Fig 2). The constraints on the parameters in our model, described in the previous section, left 6 free parameters for formal fitting. Their ranges used for the data fit are shown in Table 2.

**Table 2.** Parameters used for the data fit and the sets of values simulated over.

| Parameter | Values considered |
| --- | --- |
| $\omega_{high}^C$ | $\{3.3, 3.4, \dots, 5\}$ |
| $\omega_{high}^G$ | $\{3.3, 3.4, \dots, 4.5\}$ |
| $\varepsilon$ | $\{0.1, 0.2, \dots, 0.5\}$ |
| $E_G(0)$ | $\{100, 110, \dots, 180\}$ |
| $\omega_{high}^\gamma$ | $\{0.042, 0.083, 0.17\}$ |
| $H_{seeded}$ | $\{1, 2, \dots, 10\}$ |

To investigate the question of how many care homes were exposed at the start of the pandemic, we ran the fitting separately for *H*_*seeded*_ fixed at 1 through 10. We simulated the model over 21,060 combinations of the remaining parameters to calculate the least-squares, for each value of *H*_*seeded*_. We investigate the distribution of the parameters in Table 2 in the top ten best fitting scenarios, for each value of *H*_*seeded*_.

### Sensitivity analysis

After identifying the parameter set that minimises the least-squares, the base case (Table 2), we performed a sensitivity analysis. We measured the change in each population’s deaths when shifting individual parameters in Table 3 from the base case. This allowed us to assess the relative impact of individual parameters on each population. We also measured the change in deaths in each population when changing pairs of the time-share parameters 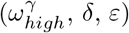, keeping all other parameters at the base case. The results were stored in a 50×50 grid and visualised using heat-maps to determine the key modes of COVID-19 ingress into and throughout care homes.

**Table 3.** Parameters involved in the sensitivity analysis. Sensitivity shift is the unit of change used for each parameter from its base case. These values were chosen to measure the change in each population’s deaths to small perturbations of individual parameters from it’s base case.

| Parameter | Sensitivity shift |
| --- | --- |
| $\omega_{end}^C$ | 1 |
| $\omega_{high}^C$ | 0.1 |
| $\omega_{low}^C$ | 0.1 |
| $\omega_{rate}^C$ | 0.1 |
| $\omega_{end}^G$ | 1 |
| $\omega_{high}^G$ | 0.1 |
| $\omega_{low}^G$ | 0.1 |
| $\omega_{rate}^G$ | 0.1 |
| $\delta$ | 0.1 |
| $\varepsilon$ | 0.05 |
| $\omega_{high}^\gamma$ | 0.0167 |
| $E_G(0) = I_G(0) + A_G(0)$ | 10 |
| $\lambda$ | 0.3 |
| $\tau$ | 0.4 |

## Results

In this section we first show how the model captures the NHS Lothian data for cases and deaths in the period from March to June 2020, and subsequently show how sensitive the results are to changes in key parameters.

### Data fit

The model captures the key features of the COVID-19 related cases and deaths in both care home and general populations, Fig 2. The minimum aggregate least-squares error was 33,042, with our model predicting 3,165 total cases and 817 total deaths compared to the total 3,123 cases and 709 deaths in the data. The average difference between data and predictions was 3.5 cases/deaths per week. In care homes we predicted 871 cases and 411 deaths compared to 903 cases and 423 deaths in the data. Our model does not predict the initial jump in deaths in care homes due to our assumption that infection reporting is constant. Further, our model overestimates the number of deaths for all populations despite a good fit for the cases, as the calculation of death rates is tied to the reporting rates.

The reproduction rates change rapidly over the period of April - May 2020, Fig 3 reflecting the delayed effect of the lockdown. The care home resident population’s fall in reproduction rate is delayed by ∼ 3 weeks compared to the fall in the general population. This delay is informed by the data, due to the *ω*_*end*_ parameters controlling the timing of the peaks in Fig 2 - we must be careful when attempting to interpret this delay.

**Fig 3.**
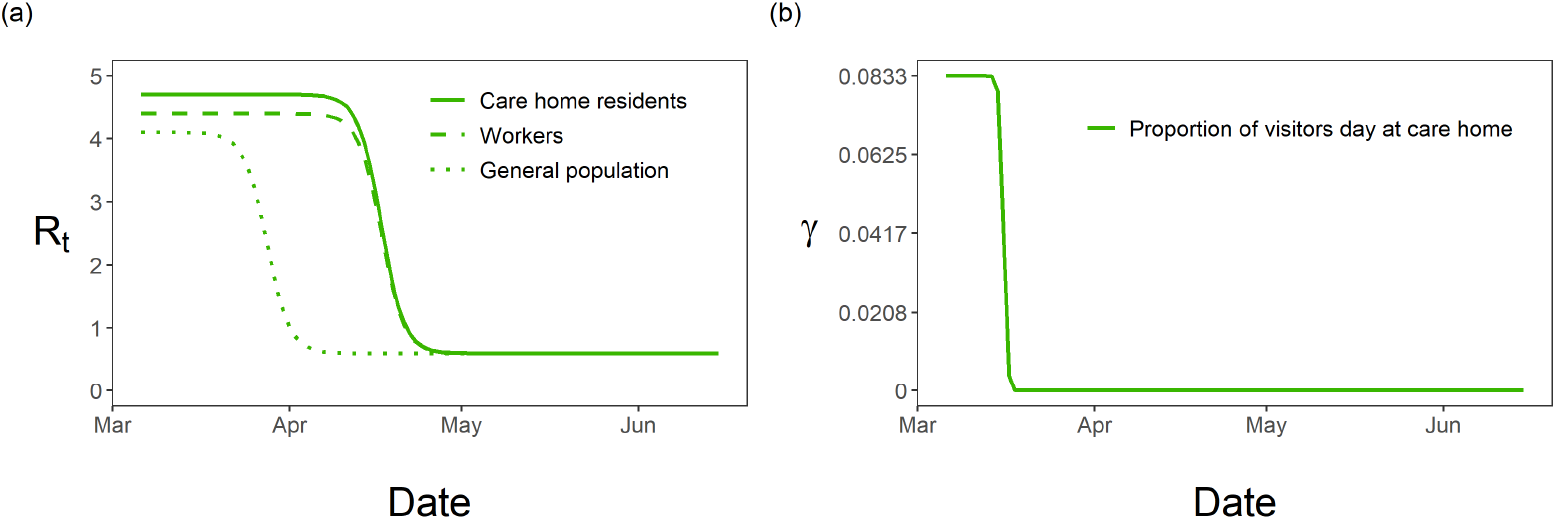
Fitted time-dependent parameters. (a) Fitted reproductive numbers over time for care home residents, *R*_*C*_(*t*), workers, *R*_*W*_(*t*), and general population, *R*_*G*_(*t*); (b) fitted visitation, *γ*, over time with drop highlighting the change in policy.

In order to assess the initial level of care home exposure to virus, we consider the quality of fit as a function of *H*_*seeded*_. The minimum sum of squares of residuals takes the shape of a parabola, with a minimum at *H*_*seeded*_ = 4, see Fig 4. This suggests that a relatively small number of homes were initially exposed to COVID-19.

**Fig 4.**
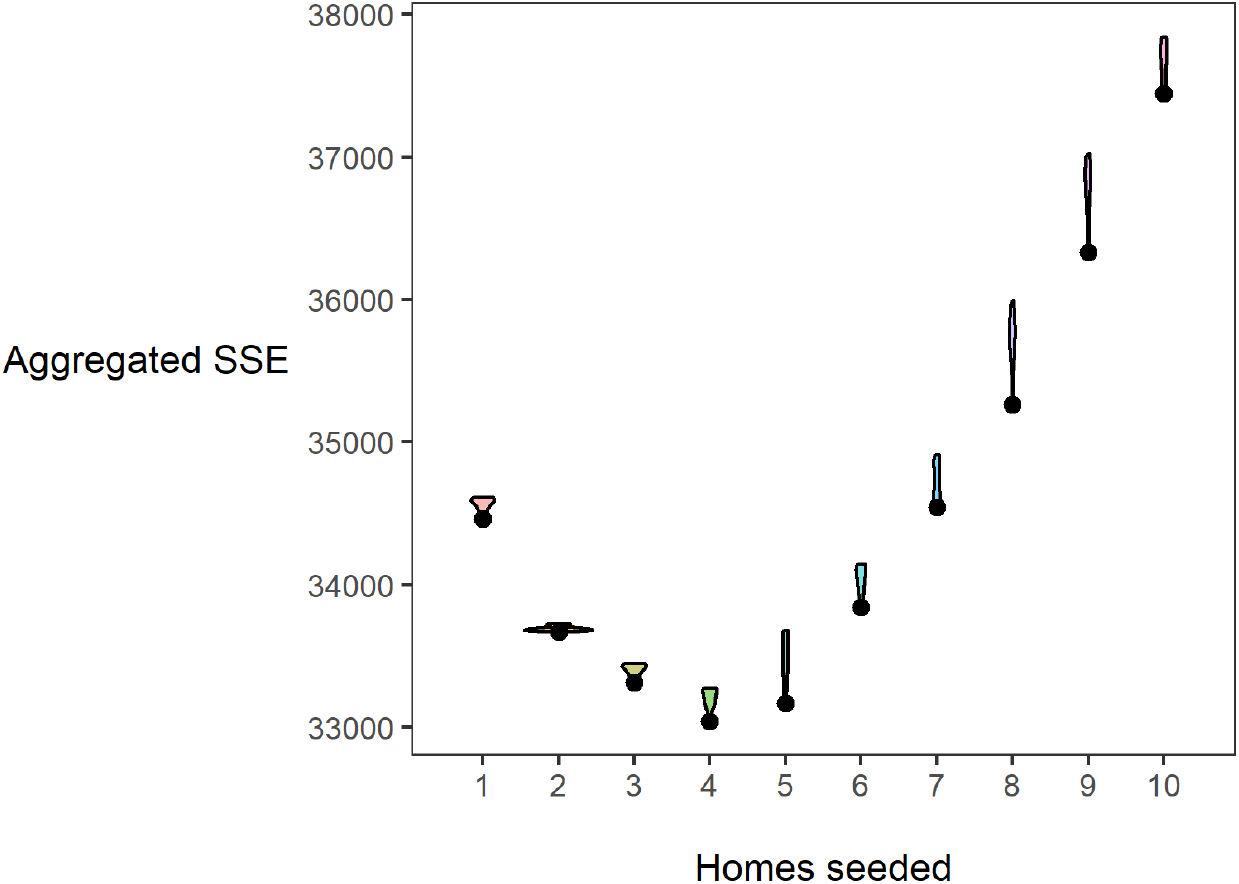
Quality of fit as a function of homes seeded. Each violin is the distribution of aggregated sum of squared errors (SSE) in the top ten best-fitting parameter sets, for a number of homes seeded. Black dots indicate the minimum aggregated SSE achieved for each home seeded.

The optimal choice (in terms of the least-squares criterion) for the parameters as used in the data fit (Table 2) is relatively stable with respect to changes in *H*_*seeded*_, Fig 5. The pre-lockdown reproduction rate in care homes, 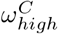, appears stable in the range of 4.5 to 4.7, changing for 10 homes seeded with the optimal value lowering to 3.9. This highlights the clear link between the reproduction rate and the exposure of care homes at the beginning of the pandemic. For the pre-lockdown reproduction rate in the general population, 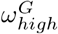, we see a stable optimal value in the range of 3.9 to 4.2. We see a lot of uncertainty in the proportion of staff shared *ε*, for *H*_*seeded*_ outside of the range of 4 to 8. The distribution for *ε* changes for *H*_*seeded*_ = 10, with the optimal value going back up to 0.5. This coincides with the rise in *E*_*G*_(0) and the substantial fall in 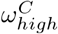. This points to the correlation between these 3 parameters. Similarly, there is a lot of uncertainty in the value of pre-lockdown visitation, 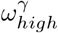, with an optimal choice for every value considered in our fitting as we vary *H*_*seeded*_. This uncertainty highlights that the parameters in our model are highly correlated. As seen in Fig 6 and Fig 7, Fig 5 can also be seen to hint at how effective these parameters are at affecting the outcome of the model. For example, the variability in the chosen value of *ε* and 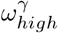 can also be attributed to the relatively small affect they have on the model outcome.

**Fig 5.**
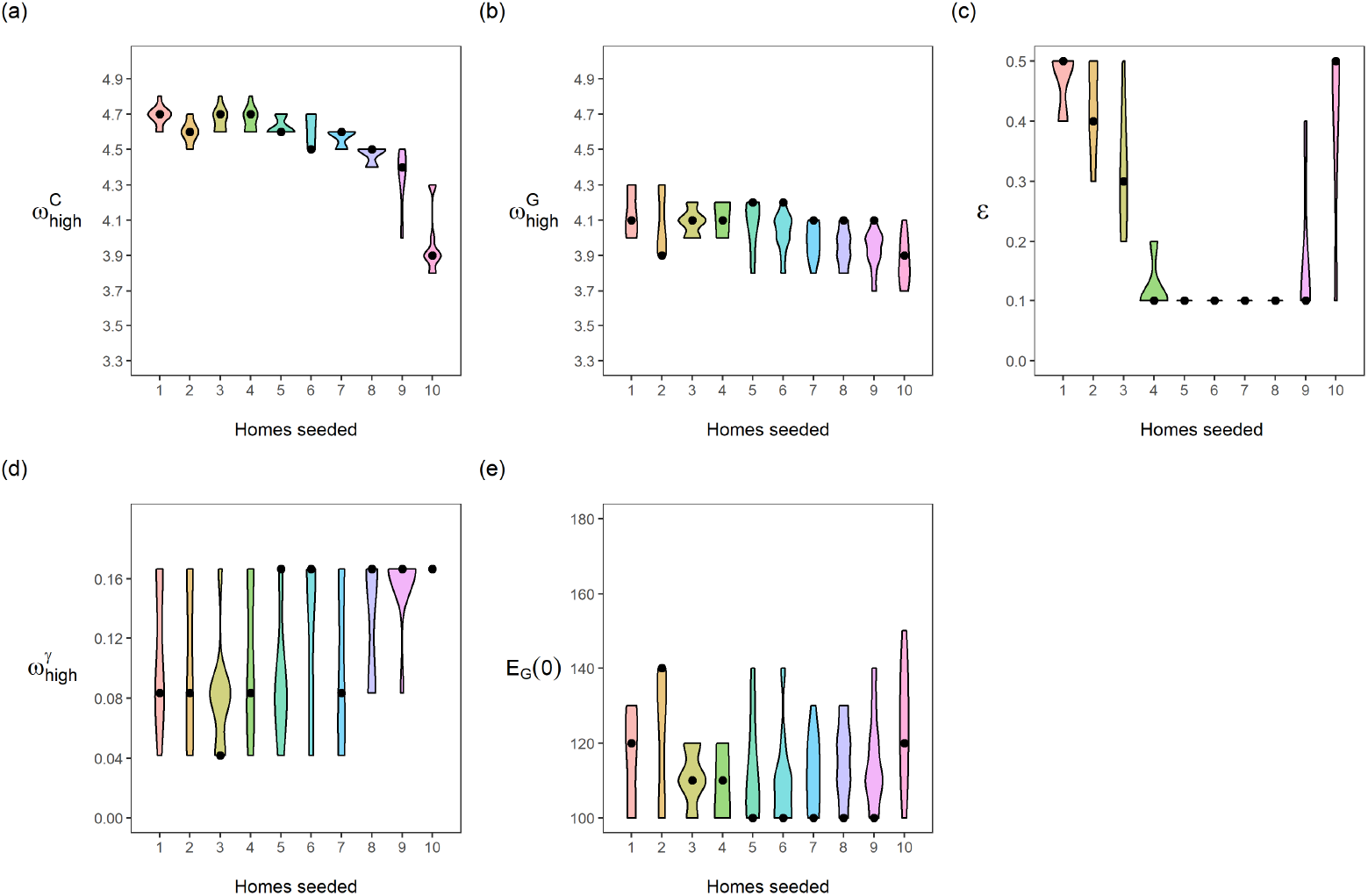
Distribution of fitted parameters as a function of homes seeded. Each panel is a different calibrated parameter. Each violin in a panel is the distribution of individual parameters in the top ten best fitting parameter sets, for a number of homes seeded. (a) pre-lockdown care home resident *R*_*t*_, 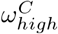; (b) pre-lockdown general public *R*_*t*_, 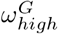; (c) staff sharing, *ε*; (d) visitation pre-lockdown, 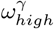 ; (e) general public seeded cases, *E*_*G*_(0) = *I*_*G*_(0) + *A*_*G*_(0). Black dots indicate the parameter value giving the lowest aggregated least square error, for each number of homes seeded.

**Fig 6.**
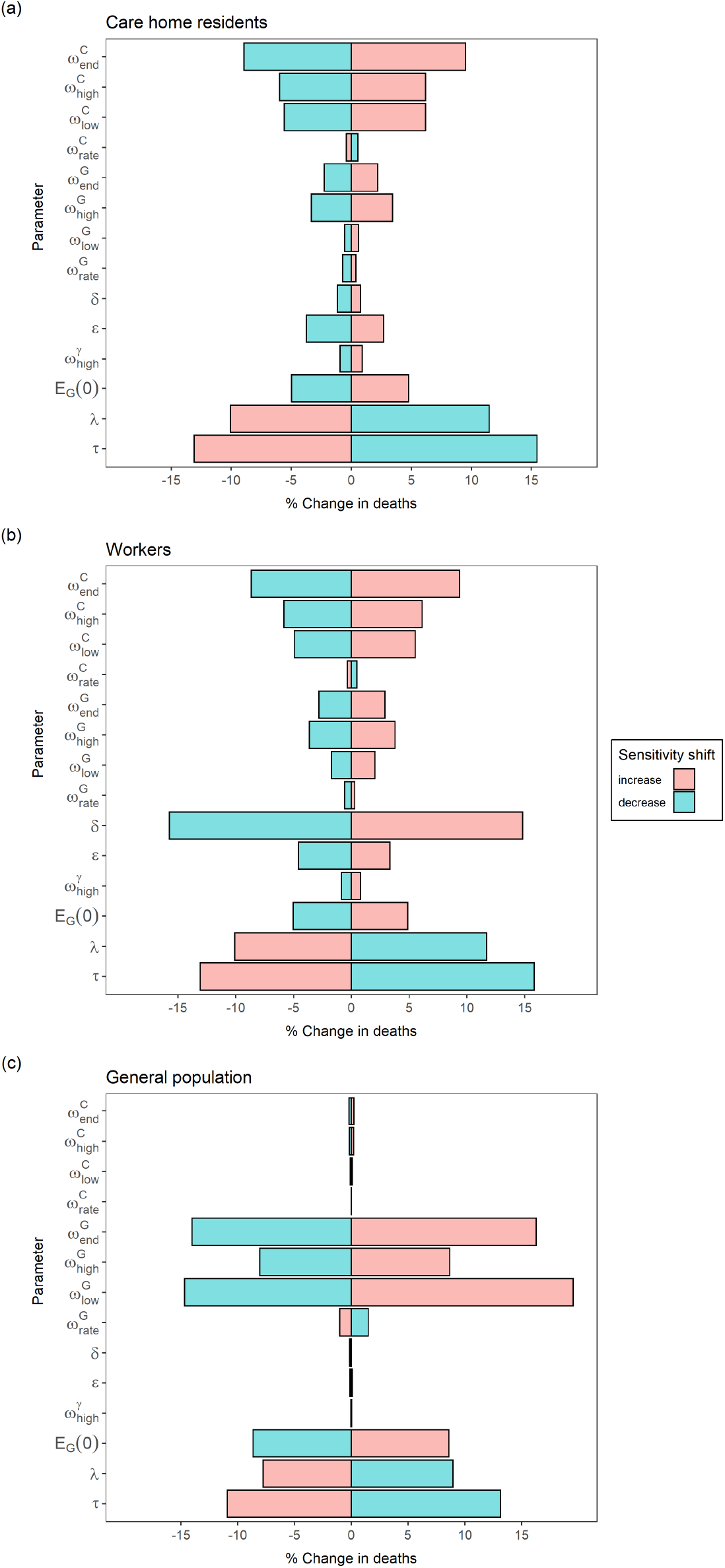
Sensitivity of the final deaths in each population to perturbations in model parameters. Each bar shows the % change in final deaths in a population caused by shifting an individual parameter from the base case, keeping all other parameters fixed at the base case (Table 1). Each parameter is increased or decreased from its base case value by the corresponding ‘sensitivity shift’ value in Table 3.

**Fig 7.**
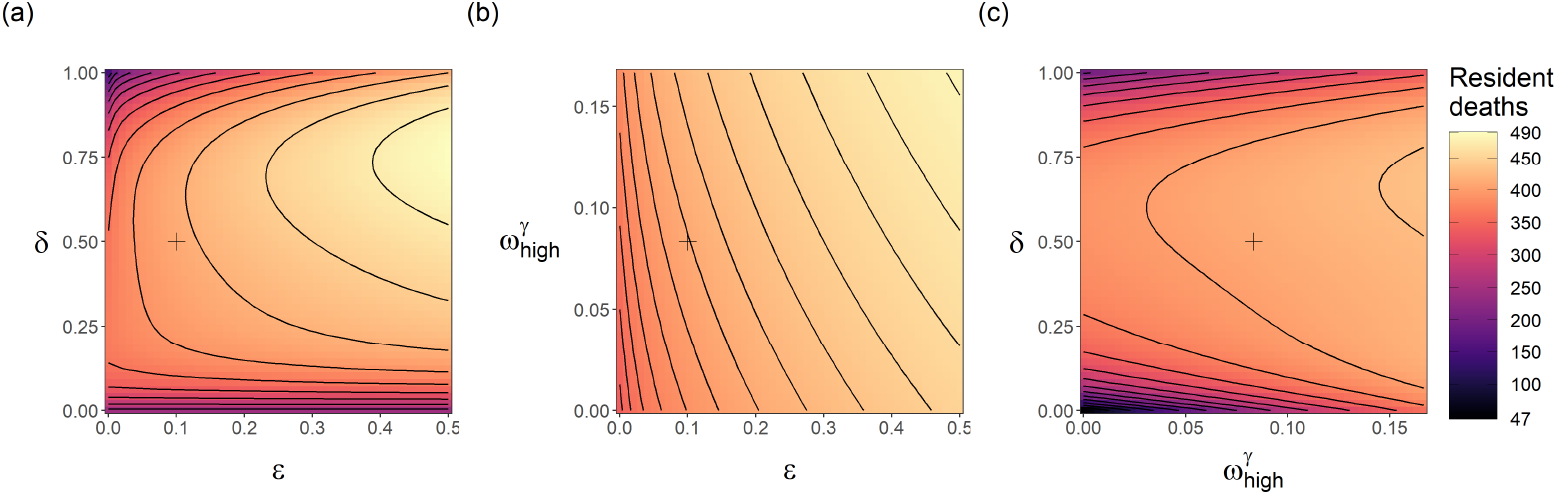
Sensitivity of the final resident deaths to the time-share/mixing parameters (*δ, ε*,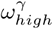). Proportion of CH staff at work is *δ*, proportion of staff shared between homes is *ε*, and pre-lockdown visitation is 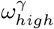. Each panel shows the combined impact of varying two of the time-share/mixing parameters, with all other model parameters fixed as the base case (Table 2). The black lines in each panel are isoclines. The cross in each panel indicates the base case value for each parameter.

**Fig 8.**
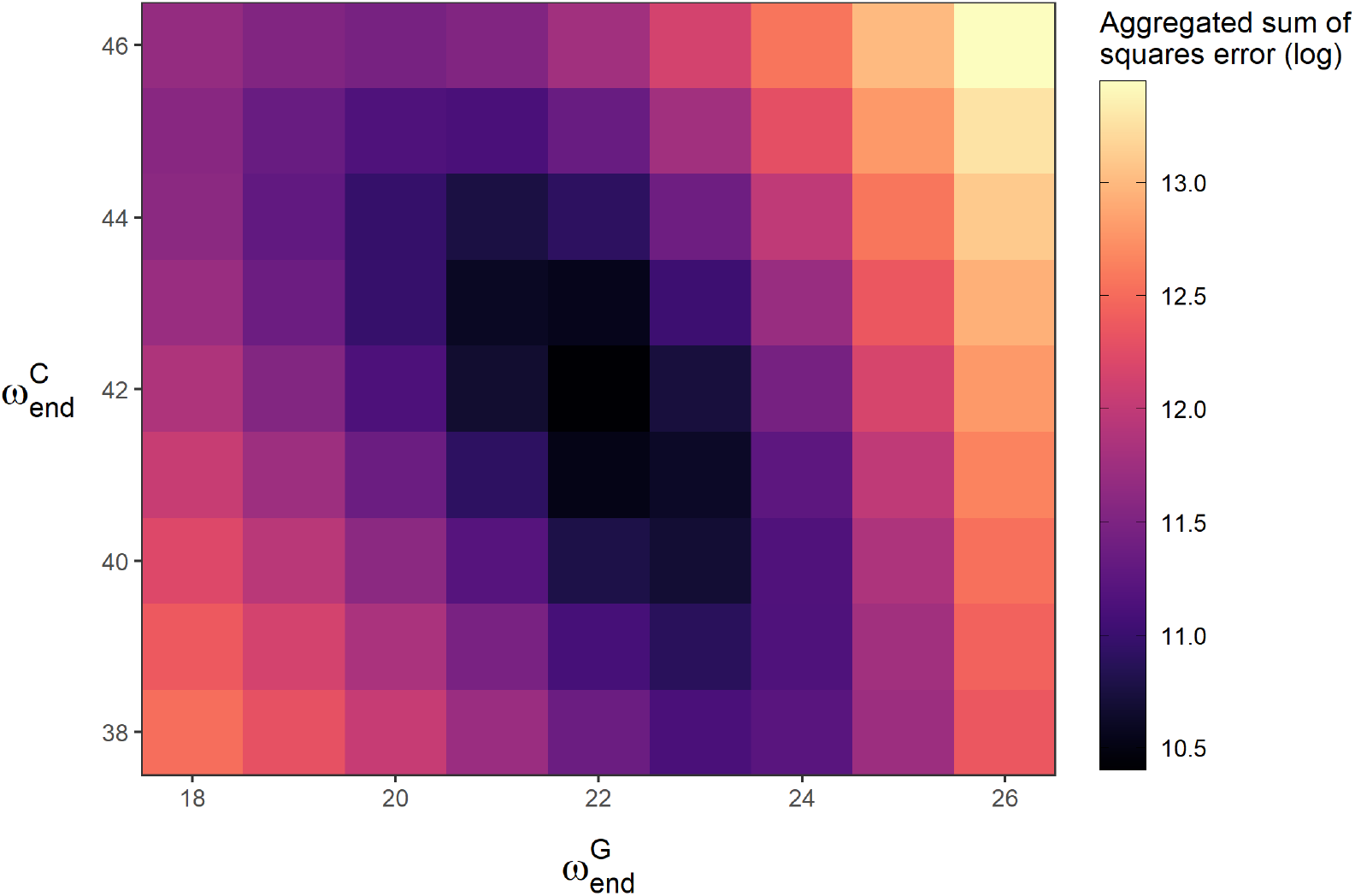
Natural log of aggregated sum of squared error in a 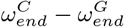 parameter space. 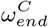 controls the timing of the drop in resident *R*_*t*_, and 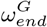 controls the timing of the drop in general population *R*_*t*_. These parameters were fitted manually, achieving a minimum for the values shown in Table 1. In the plot, all other parameters are held at the base case (Table 1).

### Sensitivity analysis

Fig 6 indicates the sensitivity of the predicted deaths to the parameters in Table 3 for care home residents, workers and general population. Predictions are most sensitive to the infectious period, *τ*, and latency period, *λ*. The parameters 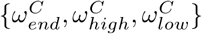 significantly influence care home and worker deaths, without affecting predicted deaths in the general population. Interestingly, a change in one day from when the care home reproduction rate drops, results in almost 10% change in predicted resident and worker deaths. The parameters, 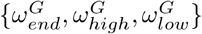, controlling the timing of the reproduction rate and it’s value before and after lockdown for the general population, significantly affect predicted deaths in the general population. Interestingly, changing the value of *δ* by 0.1 (20%) results in very little effect to the residents (*<* 5%) and general population (*<* 1%) but a 15% change in predicted worker deaths.

In our model, the path of staff catching infections from the community is controlled by *δ*; staff spreading infections between homes through staff sharing by *ε*; and visitors bringing infections into homes from the outside community by 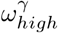. We investigate the relative risk of these COVID-19 pathways into care homes using Fig 7. This shows the combined impact of varying pairs of these parameters on the total resident deaths. Changing staff sharing, *ε*, and staff shift patterns, *δ*, the final number of predicted resident deaths do not change significantly, apart from when *δ* ∼1 (Fig 7). When considering *δ* = 1, we should also restrict *ε* = 0, as staff living in care homes would not be shared across them. Varying *ε* and 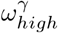 does not significantly effect predicted deaths while holding all other parameters constant at their respective values in Table 1. Changing staff shift patterns (*δ*), has the largest impact on the predicted deaths, especially in the extremes. Reducing *ε* and 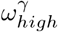 alone are weaker, but *ε* has a comparatively larger effect than 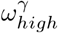. Reducing *ε* to 0 and increasing *δ* to 1 together creates an even higher impact on the predicted deaths Fig 7, showing an outbreak severity reduction of ∼ 75% compared to the observed values in Lothian during the first wave. Reducing pre-lockdown visitation from 2 visiting hours per resident to 0 hours per resident, would reduce our predicted first-wave deaths by 20 to 40, about 10% of the death count in the first wave in NHS Lothian.

## Discussion

To identify and rank the key modes of ingress into care homes, we used a combination of modelling, data fit and simulations. We find that homes in our model are more at risk to outbreaks through staff infections from the general population, relative to visitation or staff sharing. We also find that outbreaks in our model were not significantly driven by hospital discharges. These findings coincide with the results from Rosello et al., who used a stochastic compartmental model on single care homes in England [9]. We additionally find that in our model the drop in within-home reproduction rate was 3 weeks behind that of the general population.

Changing worker shift patterns in care homes (*δ*) only weakly affects the model outcome for most “reasonable” values e.g. a 2-shift pattern (*δ* = 0.5) or 3-shift pattern (*δ* = 0.33). It is only in the extremes where substantial differences are seen. *δ* close to 1, alongside low pre-lockdown visitation, greatly reduces the final outbreak size in care homes. Thus, our results point to a strategy of staff living-in with residents, in conjunction with timely lowering of visitation, as an effective pandemic response. This happened in France, where care home outbreak size was reduced significantly in care homes where staff self-confined [36]. If living within the care home is not possible, this result of very high levels of *δ* may imply that the strategy of segmenting the staff away from both care home residents and the general population whilst they are not at work would be effective, e.g., organisation of accommodation for care home workers [3]. From our model, we see the most effective solution to keeping care homes safe from infection is to focus on the pathway from general population to workers to residents.

Eliminating staff sharing did not eliminate outbreaks in our model simulations, suggesting that this was not the primary route of infection entering homes. Supporting the literature [3, 20, 37], reducing staff sharing does reduce the outbreak severity; in our model, this impact is low compared to other routes. This conclusion is limited due to our assumption of the circle sharing contact structure, which in turn reflects limited data availability regarding the contact structure of the care home industry in Lothian (due to commercial sensitivity). A different contact structure could result in staff sharing leading to more/less exportation of infection from homes with outbreaks. A more thorough examination on the contact-structure of this system and how that impacts disease spread dynamics would be an important contribution to the literature. One way to achieve this would be to consider an addition of highly connected hubs [38]. We expect that including highly connected worker populations will increase the effect of staff sharing. In our simulations, staff sharing has an effect when there exists a non-uniform distribution of infections in worker sub-populations. Worker sub-populations acting as hubs would acquire disease quicker and skew the distribution of infection amongst worker sub-populations. However, the general population strongly connects all nodes in the network, and dominates the impact of the staff sharing network on disease spread. This is because we assume a single general population with full mixing. At the geographical scale we model (a health board) this is an appropriate assumption, although it would not hold for larger scales of heterogeneity, e.g., the national level (Scotland).

A reduction in visitation reduces predicted resident deaths, as speculated in [3]. Our model predictions support findings that visitation to care homes was not the driving cause of infection in care homes [39]. Since visitation was banned, the evidence for visitation causing outbreaks is limited. Investigation with constant visitation would be necessary to see how the outcome would be different if visitation did not change at all; this was not the focus of our investigation.

Our parameter estimations suggest that after the nation-wide lock down, *R*_*t*_ within care homes dropped three weeks after it did in the wider population. Several possible explanations exist, including differences in testing availability or testing strategies and the difficulty in controlling care home resident interactions to lower disease transmission. Dropping care homes’ reproduction rate 1 day earlier, results in a 10% reduction in resident and worker deaths. Therefore a more thorough investigation into the delay between the general populations’ and care homes’ reproduction rate is necessary to aid closing this gap in future outbreaks.

From the data fit, there were a low number of care homes infected at the beginning of the first wave (*H*_*seeded*_ = 4). These initial infections could represent hospital discharges or any other pathway. The result supports the claim from a report on English care homes that resident discharges from hospitals were not the primary cause of care home outbreaks [40]. The hospital discharges are only included as initial care home infections, *H*_*seeded*_. In reality, discharges continued during the first wave [14] and more detailed data will be needed to address this problem.

We made a number of simplifying assumptions. Our model does not explicitly account for the variation in susceptibility with age [41]. It is only implicitly addressed by considering different values of *β* within and outwith of care homes, while keeping the staff and general population homogeneous. Due to the unavailability of data regarding care home worker infections, we expressed worker transmission rates in terms of transmission rates for care home residents and the general population. We assumed the resident-resident and resident-worker transmission rates were equal. However, contacts between care giving staff and residents are likely more frequent and closer than between residents. On the other hand there may be more adherence or better knowledge of how to use PPE among staff. Also, contact between residents could be reduced more easily during the pandemic [3].

The data fit was achieved by minimising the aggregated sum of squared error for each of the four time-series. This method requires the errors to be independent, follow a normal distribution and for the variance to be constant. With the data, we could not estimate the variance over time. To mitigate the effect of the differing variance of the data sets, we shifted the four time-series to the same scale, this being weekly cases/deaths.

The size of individual care homes is believed to be the main factor that influences the likelihood of a care home outbreak [14, 20, 42]. Larger homes typically have more staff and therefore a higher chance of experiencing an outbreak before the smaller ones. In general, we expect larger care homes to receive an increased force of infection from all sources, proportional to its increased size, and therefore an increased outbreak risk. This in turn could increase risk for smaller homes directly connected to the larger ones through staff sharing and visitations, and the overall outbreak risk. However, this effect could balanced by a lowered risk associated with small care homes, with the total population size kept constant. In our model, we assume a uniform home size in order to keep the model generic. As a result (and since the model is deterministic) the risk of staff and visitors bringing in infections is the same for all care homes, which may result in underestimation of the initial rate of spread. An obvious extension of the paper would be to consider various sources of heterogeneity, including size.

The National Records Scotland death data used were the dates of death registration, not the date of death. This is limiting, as we are an average of 3 days behind in the prediction of deaths [25]. The data for care home resident deaths includes deaths in hospital; including nosocomial infections, which we do not take into account into our model. We expect this not to limit the interpretation of our results, considering hospital deaths of care home residents make up approximately 5% of the total care home resident deaths [25].

We do not distinguish explicitly between symptomatic and asymptomatic individuals, as in (**A**) there are people with symptoms that would have been missed by testing. However, asymptomatic infections implicitly affect this model’s reporting rates. We do not explicitly model self-isolation or any behavioural change after infection, nor delays or changes in reporting. For simplicity, reported infected individuals are reported immediately. Since we are not explicitly modelling behaviour change once infected, we do not expect incorporating reporting delays to significantly affect our results on the infection pathways. Reporting differed over time, especially in the early weeks of the pandemic when testing was scarce; in care homes, the national policy was to test only the first few symptomatic residents [14]. With our constant reporting rate, we overestimate the number of positive cases prior to the policy change, and underestimate the cases afterwards. A time-dependent reporting rate would impact our death rate and reproductive rate parameters and therefore requires further study.

Data regarding care home outbreaks were limited due to the commercial nature of care home organisations in Scotland. Making this data available would allow for additional modelling approaches. Adding in further heterogeneity into the system by including a distribution of home sizes and types would further improve the modelling approach. Including a stochastic component to this model could lead to more insight into “super-spreader” events in care homes [35] and their effect on epidemic response. Finally, this work focuses on disease dynamics over the course of the first wave, it would be interesting to model this system for future waves and to incorporate the implementation of vaccinations. Extending this system to the rest of Scotland or even the entirety of the UK would be a clear extension of this work.

## Conclusion

In our model, the primary route of introduction to the care home sector is from the staff via the general population. Our results also agree with other findings in the literature which suggest that visitation was not the major cause of COVID-19 establishment in care homes [39] and that hospital discharges accounted for a small number of care home outbreaks early on in the pandemic [40]. Prioritising the protection and monitoring of staff, second to timely reductions in visitation and staff sharing, are what our model suggests for an effective reduction in outbreak size. Our findings also show difficulties in halting the spread of COVID-19 within the homes in tandem with the general population. This highlights the need for more planning and support for care homes and their staff in organising quick and effective responses to emerging pandemics. The lessons to be learned from the first wave of COVID-19 will be crucial to minimise the damage from future outbreaks.

## Data Availability

The publicly available data used has been referenced and hence can be found there.
The care home COVID-19 case and death data can be requested from Prof. Bruce Guthrie (https://doi.org/10.1016/S2666-7568(20)30012-X).
All code used for the results in this manuscript are being stored on GitHub at https://github.com/ewanmct/COVID-care-homes

## Acknowledgements

The authors thank Public Health Scotland for their support, and Prof. Bruce Guthrie (University of Edinburgh) for sharing the Lothian care home data. EMT has been partially supported by the University of Strathclyde Student Excellence Award and MB by Defra. Results were obtained using the ARCHIE-WeSt High Performance Computer (www.archie-west.ac.uk) based at the University of Strathclyde. Results were also obtained using the Pin cluster at the University of Stirling.

## Appendices

